# Neural emotion regulation during pregnancy – a fMRI study investigating a transdiagnostic mental health factor in healthy first-time pregnant women

**DOI:** 10.1101/2024.09.13.24313410

**Authors:** Franziska Weinmar, Lydia Kogler, Elisa Rehbein, Carmen Morawetz, Inger Sundström-Poromaa, Alkistis Skalkidou, Birgit Derntl

## Abstract

Pregnancy is a psycho-neuro-endocrinological transition phase in which a plethora of hormone levels rise substantially, modulating socioemotional functions, brain structures, and networks and thus presenting a window of vulnerability for mental health. A transdiagnostic factor for psychopathology is emotion regulation, which is influenced by sex hormones, such as estradiol (E2), across the menstrual cycle on the behavioral and neural level. Whether this is also the case in the antepartum period remains unknown. For the first time, behavioral and neural emotion regulation were investigated in healthy primiparous pregnant females with extremely high E2 levels during the second trimester (N = 15) using a functional magnetic resonance imaging (fMRI) paradigm. Results were compared with naturally-cycling females with high E2 levels (after E2-administration, N = 16) and low E2 levels (early follicular phase, N = 16). Although pregnant females reported the lowest trait use of cognitive reappraisal, all females successfully regulated their emotions by applying cognitive reappraisal in the scanner. On the neural level, all females had increased activity in the left middle frontal gyrus during downregulation of negative emotions. Pregnant females showed no significant differences in functional connectivity (psychophysiological interaction, resting-state) related to emotion regulation compared to the nonpregnant group. However, group differences emerged for amygdala activation. In pregnant females, increased amygdala activity predicted reduced regulation success and was positively associated with depression scores. This first fMRI study during pregnancy indicates that depression scores are reflected in heightened amygdala activity already observable in the antepartum period. Thus, through its association with reduced regulation success, increased amygdala activity suggests a neural risk marker for peripartum mental health. Future research needs to investigate emotion regulation in pregnant and postpartum women to further understand pregnancy-related changes and associations of mood, emotional and neural functions. Eventually, this will allow enhanced identification, prevention, and treatment of peri- and postpartum mental ill-health.

## Introduction

Pregnancy is a physiological and psychosocial transition phase presenting a window of vulnerability for mental health. Anxiety and depression rates increase up to 15% during pregnancy and after giving birth, 10-20% of women suffer from postpartum depression (PPD; 1-3). Sex hormone fluctuations have been associated with the onset of mental disorder symptoms and are suggested to play a role in the pathophysiology of pregnancy-related mood disorders (3–6). During pregnancy there is a drastic surge in sex hormones including estradiol (E2), exceeding any levels females will experience in their entire non-pregnant life (5,7). In the postpartum period, E2 levels drop sharply, returning to beyond pre-pregnancy levels in only about five days (3,5,8). By passing the blood-brain barrier due to its lipophilic properties along with widespread expression of E2-receptors, E2 can influence the brain (6,9). Considering the incomparable rise and fall of E2 levels during and following pregnancy, neuroimaging studies have confirmed neuronal plasticity and neural network reorganization (for reviews see 3,10,11). Affected areas are associated with social cognition and emotion processing and overlap with the Theory-of-Mind- and the Default Mode Network (DMN; 12-14). Recent data on pre-vs post-pregnancy resting-state functional magnetic resonance imaging (fMRI) demonstrated an increase in temporal coherence in the DMN (14), which promotes the assumption that neural structural and connectivity changes during pregnancy might serve a behavioral adaptation purpose in the transition to motherhood (12–14). Even though only few studies have examined emotional functions during pregnancy, changes are evident (15–18). In terms of self-reports, pregnant females indicate higher mood instability and emotional sensitivity, as well as reduced emotion regulation, which were related to sex hormone levels (2,16,19). Further, studies have shown that pregnant females have improved accuracy for encoding threatening facial expressions and an enhanced sensitivity to negative stimuli (2,16,18). This may be explained by the effect of enhanced hormones, mainly E2, on fronto-amygdala circuits (2). In the postpartum phase, a positive correlation of amygdala response to emotional stimuli and PPD symptoms was demonstrated, suggesting enhanced arousal for salient stimuli in mothers with PPD (20). However, research on behavioral and neural emotional functions *during pregnancy* is still limited, even more so regarding emotion regulation (3,5).

The ability to regulate emotions is a requirement for intact social interaction and fundamental for our well-being and health (21–23). As impairments in emotion regulation are present in many disorders and contribute to the development and/or maintenance of psychopathology, emotion regulation is proposed as a *transdiagnostic factor* for mental health (21,23–25). The most frequently studied strategy to regulate emotions is *cognitive reappraisal*, which targets the process of cognitive re-evaluation (21,22). General and distinct neural networks are assumed to underly emotion regulation and the different strategies, most of them including the bilateral inferior frontal gyrus (IFG) and middle frontal gyrus (MFG; for meta-analyses see 26-28). Furthermore, fronto-amygdala coupling seems particularly relevant for successful emotion regulation (29). A recent meta-analysis on task-dependent functional connectivity during emotion regulation reported task-modulated coupling between prefrontal regions and the left amygdala, which could be enhanced with regulation success (30). Here, specifically the *left* IFG seems to play a key role and a direct link between the *left* IFG and the *left* amygdala is suggested during emotion regulation (27,30,31). In contrast, deficits in emotion regulation might be explained by a failure to adequately recruit the neural regulation networks (25). Modulating factors of emotion regulation on the behavioral and neural level are sex hormones, especially E2 (32–34). Previous studies have shown that emotion regulation success is influenced by E2 levels throughout the female menstrual cycle: Whereas increased regulation effort is required in phases of low E2 (33,34), also reflected in enhanced recruitment of frontal neural resources (33), females are more successful at downregulating emotional arousal in high E2 phases (35). To date, however, little is known about emotion regulation abilities during phases of extremely high E2 levels, such as pregnancy. In pregnant women, inadequate self-reported emotion regulation has been associated with higher hair cortisol levels indicating chronic stress, sleep problems, substance use, and increased rates of depression, anxiety, self-injurious thoughts and behaviors (36–40). After birth, the use of regulation strategies was found to predict PPD symptoms, whereby women diagnosed with PPD report less frequent use of adaptive strategies (41–44). Most importantly, emotion regulation is suggested to have long-term implications for parental health, caregiving behavior as well as health and development of the child and is considered a protective factor for psychopathology of mother and child (39,40). Nevertheless, no study to date has examined behavioral and neural emotion regulation during the antepartum period, although changes in brain structures related to these functions are evident after pregnancy (13).

Addressing this gap in research, the present study investigated behavioral and neural emotion regulation for the first time in primiparous pregnant females using a standard emotion regulation paradigm for negative emotions during fMRI. Outside the scanner, behavioral responses in a positive emotion regulation paradigm were assessed, together with several self-report measures. To specifically investigate the influence of E2, results were compared to nulliparous naturally-cycling females with high and low E2 levels. Based on previous findings (19), we hypothesized that pregnant females self-report less application of emotion regulation strategies. As no previous study has examined peripartum emotion regulation beyond self-report, our hypotheses were partly exploratory on the behavioral and neural level. However, referring to evidence of altered emotional functions during pregnancy (2,16–18), we hypothesized reduced regulation success in the pregnant compared to the nulliparous groups. Given results on altered ante- and postpartum neural structure and function (12,13,17,18) and the influence of E2 on emotion regulation (33,34) we hypothesized group differences on the neural level, particularly in the amygdala (9) and frontal regions (IFG, MFG; 33,13) as well as task- and resting-state functional connectivity (14,20). Further, we hypothesized associations between emotion regulation abilities and mood symptoms. Here, we anticipated relations of regulation success with functional activity, specifically positive for frontal regions and negative for the amygdala (29,31,33,45). Additionally, we expected relations of reduced regulation success (1,41) and altered neural activity (20,46) with increased depression symptoms. As such, the present study sheds light on underlying neural mechanisms involved in emotion regulation during pregnancy, which ultimately has implications to understand specificities of emotion (dys)regulation in the peripartum phase.

## Material and Methods

### Participants

Forty-seven human females between the age of 19-36 years were included in the present study, of which 32 were naturally-cycling (NC) and 15 primiparous. All participants were right-handed, did not have any present or past mental, neurological, or endocrine disorders and did not take hormonal contraceptives during the past six months or any other medication. NC females were required to have a regular, natural menstrual cycle between 26-32 days and no current or past pregnancies. NC participants were equally randomized to the E2 valerate (E2V; N = 16) or placebo group (N = 16), receiving either an E2V or placebo pill, respectively. Pregnant females were in the second trimester of their pregnancy (21st-28th gestational week) and provided a copy of their ultrasound screening to exclude any pregnancy-related complication in the mother or fetus. Participants were recruited via the University Tübingen e-mail provider. Pregnant females were additionally informed about the study in the University Women’s Hospital Tübingen and in surrounding gynecology practices. All participants were offered 100€ for participation. Written informed consent and the data protection agreement was obtained from all participants before inclusion. The study was approved by the Ethics Committee of the Medical Faculty of the University of Tübingen (754/2017/BO1) and data were collected from 08/2018 till 12/2021.

### Procedure

The present study was part of a project on pregnancy and the brain. Some of the participants included in the current sample have previously been part of samples in other publications within this project (12,33). The procedure for the study is summarized in Figure 1a and outlined below.

**Figure 1.**
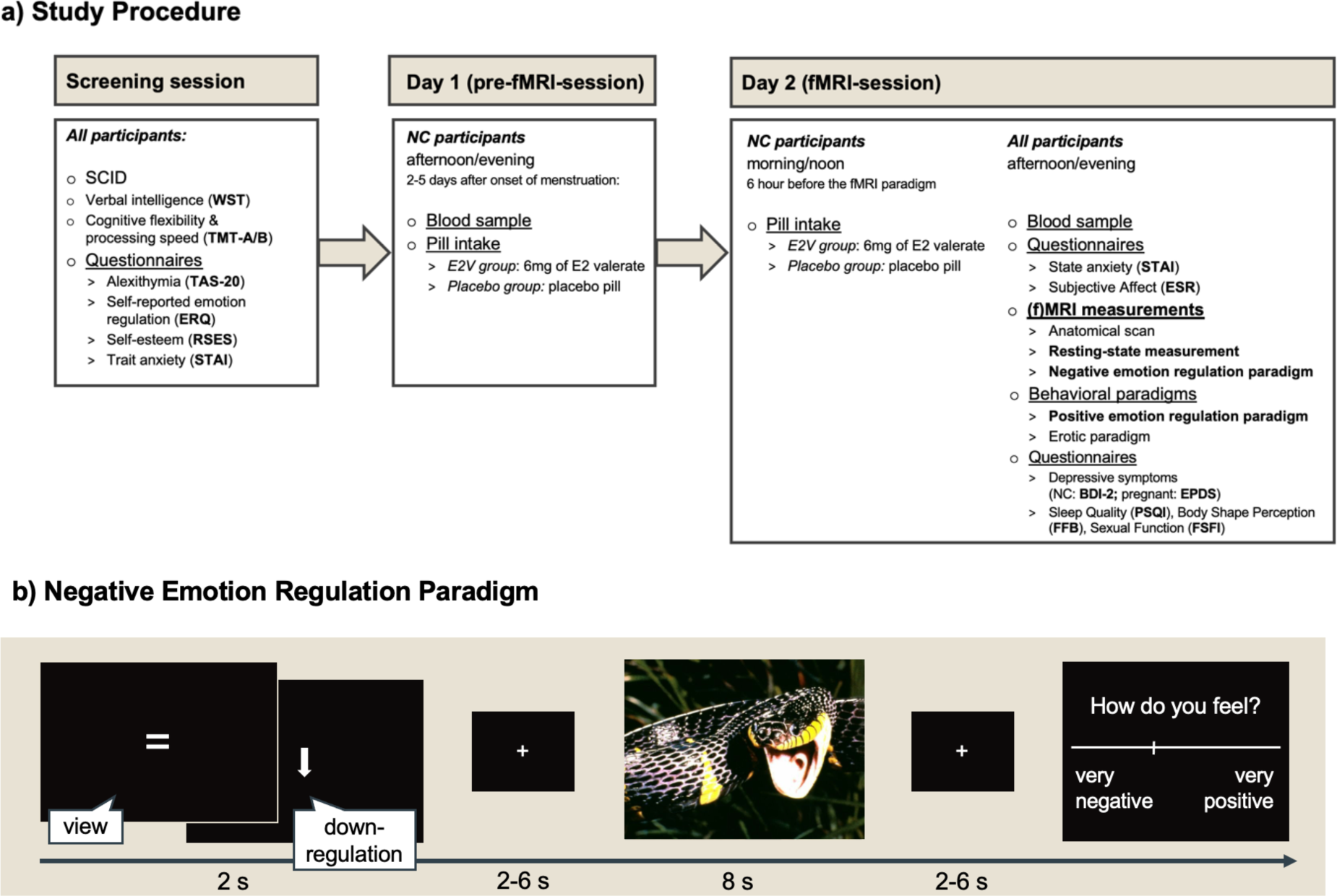
Study procedure (a) and negative emotion regulation paradigm (b) Note. a) Study procedure. All participants were screened for past or present mental disorders (SCID; 55) and cognitive abilities were assessed (verbal intelligence, WST; 56; processing speed and cognitive flexibility, TMT-A/B; 57). NC participants were randomized to receive either an E2 valerate or placebo pill 2-5 days after the onset of their menstruation and one day before the fMRI session (day 1). On the following day (day 2) NC females had a second pill intake 6h before the fMRI session to maintain high E2 levels. In the fMRI session (day 2), all females performed the negative emotion regulation paradigm while being in the MRI-scanner and the positive emotion regulation paradigm outside the scanner. The following self-report measures were assessed: alexithymia (TAS-20; 58), self-esteem (RSES; 59,60) state and trait anxiety (STAI; 61,62), subjective affect (ESR; 63), sleep quality (PSQI; 64), body shape perception (FFB; 65), sexual function (FSFI; 66), trait use of emotion regulation strategies (ERQ; 49,50) and depressive symptoms in pregnant (EPDS; 51,52) and NC females (BDI-2; 53,54). Results of measurements indicated in bold are reported in the present study. b) Negative emotion regulation paradigm. At the beginning of each trial, participants were instructed to either view (indicated by an equal sign) highly negative pictures or to downregulate their emotional response (indicated by a downward-pointing arrow). Pictures taken from the International Affective Picture System (68) were presented on black background with a size of 800×600 pixels and a visual angle of 32°x24° and were followed by a jittered fixation cross. After each picture, participants rated how they felt on a continuous scale ranging from very negative to very positive. Figure adapted from Rehbein et al. (33). Abbreviations: BDI-2, Beck-Depression-Inventory-2; E2, estradiol; E2V, estradiol valerate; EPDS, Edinburgh Postnatal Depression Scale; ERQ, Emotion Regulation Questionnaire; ESR, Emotional Self-Rating; FFB, Fragebogen zum Figurbewusstsein (Body Shape Questionnaire); FSFI, Female Sexual Function Index; NC, naturally cycling; PSQI, Pittsburgh Sleep Quality Index; RSES, Rosenberg self-esteem scale; SCID, Structured Clinical Interview; STAI, State Trait Anxiety Index; TAS-20, Toronto Alexithymia Scale; TMT-A/B, Trail making test A/B; WST, Wortschatztest (Vocabulary test).

All participants were screened for past or present mental disorders and cognitive abilities were assessed (see Figure 1a). NC females reported onset of their menstruation and were invited to the laboratory between day 2 and day 5 of their menstrual cycle, i.e., their early follicular phase. One day before the MRI assessments (*day 1*) blood was drawn in NC females and participants of the E2V group received 6 mg of estradiol valerate (Progynova® 21; Bayer Weimar GmbH & Co. KG) whereas participants in the placebo group received placebo pills in a double-blind fashion. On *day 2*, NC participants took a second dose approximately 20-24 h after the first pill intake and 6 h before the fMRI session. This procedure allows the experimental elevation of E2 levels, comparable to the peri-ovulatory phase, while at the same time maintaining low progesterone and testosterone levels (47,48). On *day 2*, all groups were invited to the laboratory, where blood samples were drawn to assess hormone levels and neuroimaging was performed. As the Ethics Committee approved only a 30 min (f)MRI session for pregnant participants, all participants performed a subsequent positive emotion regulation task outside the MRI-scanner. Additionally, several self-report measures were assessed (see Figure 1a). Trait use of emotion regulation strategies was reported on the emotion regulation questionnaire (ERQ; 49,50) and depressive symptoms were assessed with the Edinburgh Postnatal Depression Scale (EPDS; 51,52) in pregnant females and the Beck Depression Inventory (BDI-2; 53,54) in NC females. The whole procedure on *day 2* took maximum 170 min.

### Emotion regulation: stimuli and fMRI paradigm

The emotion regulation paradigm was adapted from previous studies (29,33,67). During the negative emotion regulation paradigm (see Figure 1b), participants were exposed to 24 highly negative pictures (International Affective Picture System; 68) and followed two different instructions: 1) For *downregulation*, participants were instructed to decrease their emotional response towards the picture, either by changing the perspective on or meaning of the picture, for example, by increasing their personal distance towards the picture, but not by thinking of something positive. 2) In the *view* instruction, participants were asked to experience arising emotions towards the picture without changing them. In an event-related design, each picture was shown twice for the duration of 8 sec; once preceded by the instruction (2 sec) for *downregulation* (downward-pointing arrow) and once for *view* (equal sign). Instructions and pictures were followed by a jittered fixation cross (2-6 sec). After each picture, participants rated their emotional state on a continuous visual rating scale (very negative to very positive). The positive emotion regulation paradigm outside the MRI-scanner is described in the Supplement.

### Hormonal assessments

Hormonal levels of E2, progesterone, and testosterone were analyzed at the central laboratory of the University Hospital Tübingen. 7.5 ml of blood was drawn in serum tubes and analyzed using enzyme-linked immunoassays (ELISA). Sensitivity and range of measurement for E2, progesterone, and testosterone were: E2: 43.60-11,010 pmol/l; progesterone: 0.67-190.80 nmol/l; testosterone: 0.24-52.05 nmol/l. Due to problems in drawing blood and technical problems, four blood samples (one placebo, two E2V, one pregnant) were missing.

### Statistical analyses

IBM SPSS Statistics (version 27.0) was used for all statistical analyses, if not specified otherwise. To assess group differences in sample characteristics, univariate Analysis of Variances (ANOVAs) were applied with group (placebo, E2V, pregnant) as factor. In case of significant group differences, Bonferroni-adjusted post-hoc tests were performed. If the assumption of a normal distribution was not met, nonparametric Kruskal-Wallis H tests were applied. For statistically significant results, follow-up Mann-Whitney U tests were calculated between the individual pairs of groups. For all analyses, the alpha criterion level was set to *p* ≤ .05. Effect sizes for significant differences in the ANOVAs are reported in ηp^2^. As this study includes a small but first sample of pregnant females during fMRI measurement, statistical tendencies are reported up to *p* ≤ .10 for brain activity results. In case data was missing for a participant, it was not replaced but the participant excluded from the respective analysis.

### Emotion regulation

Emotional state ratings acquired per picture and instruction were summarized in mean values for the view and regulation condition for each participant. To compare between groups and conditions, mixed between-within-subjects ANOVAs were performed with mean state ratings as a dependent interval variable, group (placebo, E2V, pregnant) as between-subjects factor, and regulation (regulation, view) as within-subject factor. To assess emotion regulation success, we subtracted mean emotional state ratings for view from mean ratings for the regulation condition for each participant (29). To compare emotion regulation success for negative pictures between groups, a univariate ANOVA was performed with group (placebo, E2V, pregnant) as factor and regulation success as dependent variable. For the negative emotional state ratings, data was missing for three females, one in each group.

### fMRI data acquisition and analysis

Data were acquired on a 3T Siemens PRISMA scanner at the University Hospital Tübingen. Brain structure was measured with a standard magnetization-prepared rapid gradient-echo sequence (MPRAGE; TR = 2.3 sec, TE = 4.16 msec, slice-thickness = 1 mm, voxel size = 1×1×1 mm, flip-angle 9°, distancing factor 50%, GRAPPA acceleration factor, sagittal orientation). The fMRI sequences (task, resting-state) consisted of standard echo-planar imaging (EPI) protocols (32 interleaved slices, TR = 2 sec, TE = 32 msec, voxel-size 3.4×3.4×3.4 mm, flip-angle 76° transversal orientation, anterior-posterior commissure orientation, 64-channel head coil). Participants were provided earplugs to reduce the sound intensity. To further ensure that the sound level was kept at a minimum for the unborn child, the volume of the MRI sequence was measured before scanning, reaching approximately 90dB. Besides, the unborn child in the womb is protected against noise disturbance through the amniotic fluids. Data from one female in the placebo group had to be excluded due to excessive head movement and technical problems and one pregnant participant terminated MRI measurements after the anatomical scan.

Data were preprocessed using SPM12. Preprocessing consisted of slice time and distortion correction, realignment and unwarping, segmentation, co-registration, normalization and smoothing (6 mm FWHM). Participants with head movement of more than 2 mm were excluded from the analyses. The first-level analysis included the regressors downregulation and view, the emotional state rating period, instruction period, plus six movement parameters and time-derivatives for each participant. At the second level, a full factorial analysis with the factors group (placebo, E2V, pregnant) and regulation (downregulation, view) was calculated. Whole-brain results were corrected for multiple comparisons with cluster-wise correction (*p* < .001). Brain areas were labeled with the Anatomy toolbox as available in SPM12. Data was visualized with SPM12.

### Task-based fMRI data

#### Region of interest (ROI) analyses

Based on previous findings (27,33) and our a-priori hypotheses, selected ROIs related to emotion regulation were bilateral IFG (MNI (x,y,z): -46, 26, -8 / 50, 30, -8) and bilateral MFG (MNI (x,y,z): -45, 20, 35 / 42, 48, -2). Additionally, due to the impact of E2 and involvement in emotion regulation, bilateral amygdala were included (MNI (x,y,z): -24, -3, -21 / 24, -6, -15). Beta-values were extracted with MarsBarR (69; 10 mm sphere around the reported MNI peaks). The effects of group (placebo, E2V, pregnant) and regulation (downregulation, view) were analyzed in a mixed between-within ANOVA. Age was included as covariate as pregnant females were significantly older than nulliparous groups (Table 1). To correct for multiple testing related to laterality, the Bonferroni-corrected significance level was set to *p* < .025.

**Table 1.**
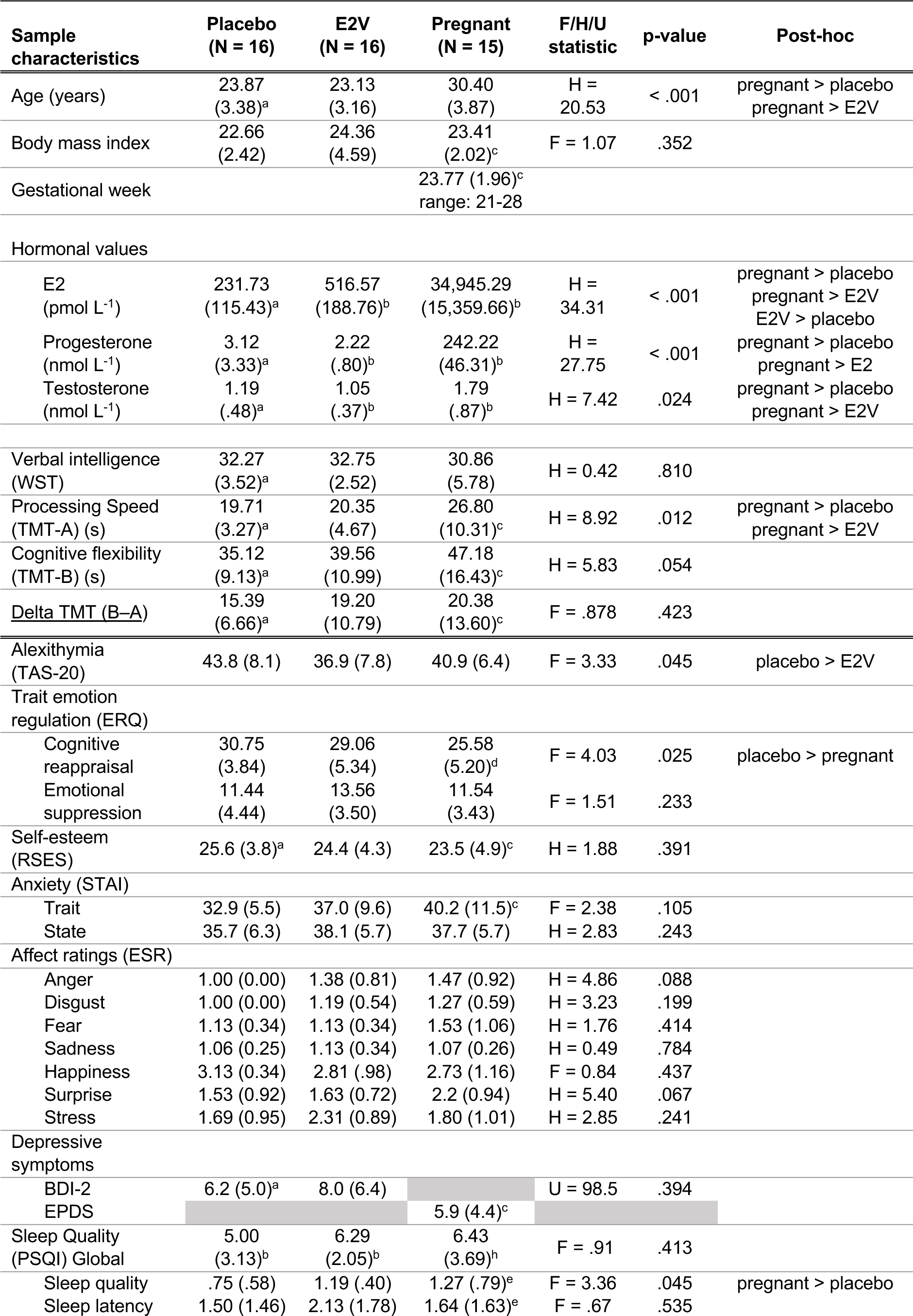
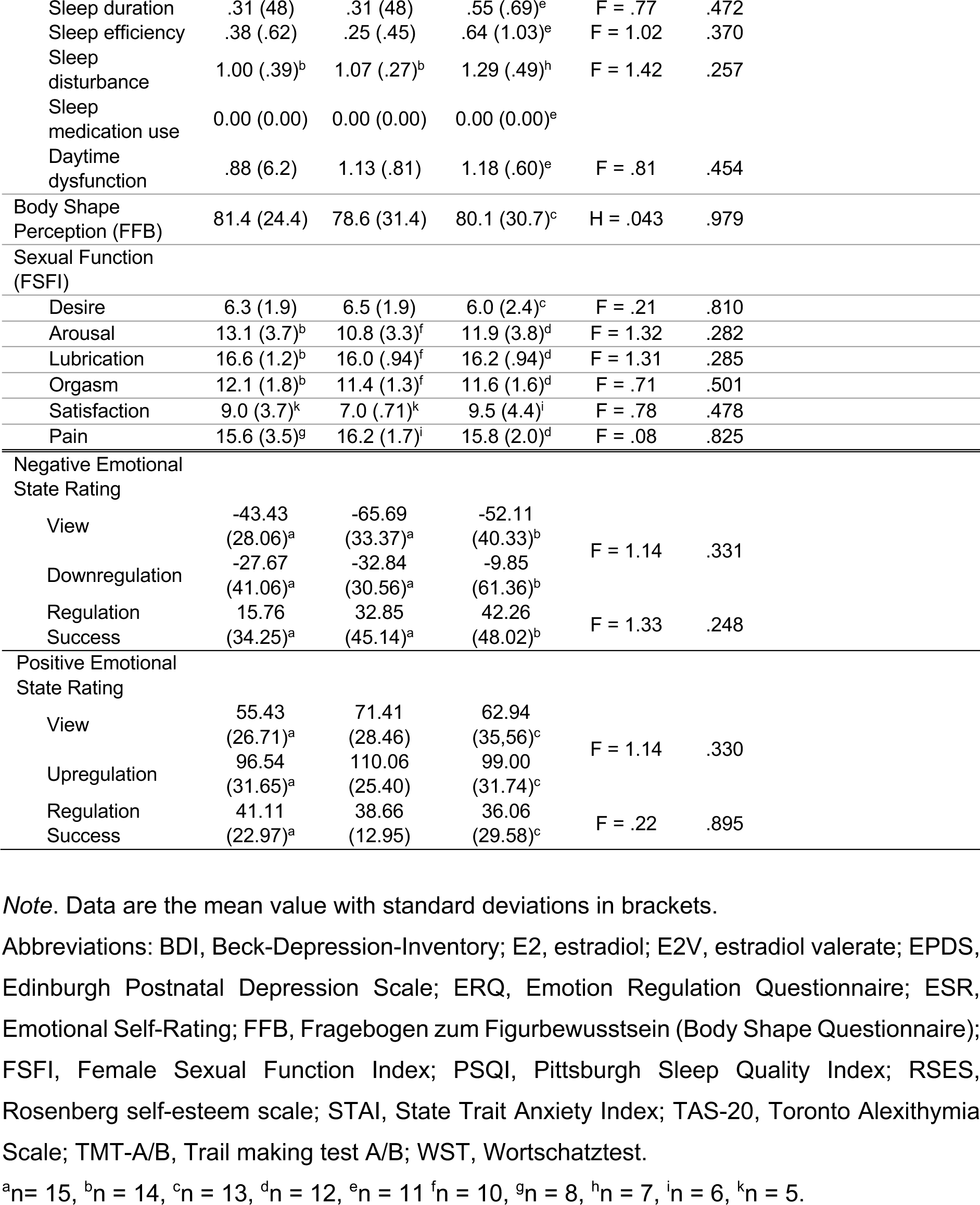
Details on sample characteristics including hormonal data and emotional state rating.

| Sample characteristics | Placebo (N = 16) | E2V (N = 16) | Pregnant (N = 15) | F/H/U statistic | p-value | Post-hoc |
| --- | --- | --- | --- | --- | --- | --- |
| Age (years) | 23.87 (3.38)<sup>a</sup> | 23.13 (3.16) | 30.40 (3.87) | H = 20.53 | < .001 | pregnant > placebo<br>pregnant > E2V |
| Body mass index | 22.66 (2.42) | 24.36 (4.59) | 23.41 (2.02)<sup>c</sup> | F = 1.07 | .352 |  |
| Gestational week |  |  | 23.77 (1.96)<sup>c</sup><br>range: 21-28 |  |  |  |
| Hormonal values |  |  |  |  |  |  |
| E2 (pmol L<sup>-1</sup>) | 231.73 (115.43)<sup>a</sup> | 516.57 (188.76)<sup>b</sup> | 34,945.29 (15,359.66)<sup>b</sup> | H = 34.31 | < .001 | pregnant > placebo<br>pregnant > E2V<br>E2V > placebo |
| Progesterone (nmol L<sup>-1</sup>) | 3.12 (3.33)<sup>a</sup> | 2.22 (.80)<sup>b</sup> | 242.22 (46.31)<sup>b</sup> | H = 27.75 | < .001 | pregnant > placebo<br>pregnant > E2 |
| Testosterone (nmol L<sup>-1</sup>) | 1.19 (.48)<sup>a</sup> | 1.05 (.37)<sup>b</sup> | 1.79 (.87)<sup>b</sup> | H = 7.42 | .024 | pregnant > placebo<br>pregnant > E2V |
| Verbal intelligence (WST) | 32.27 (3.52)<sup>a</sup> | 32.75 (2.52) | 30.86 (5.78) | H = 0.42 | .810 |  |
| Processing Speed (TMT-A) (s) | 19.71 (3.27)<sup>a</sup> | 20.35 (4.67) | 26.80 (10.31)<sup>c</sup> | H = 8.92 | .012 | pregnant > placebo<br>pregnant > E2V |
| Cognitive flexibility (TMT-B) (s) | 35.12 (9.13)<sup>a</sup> | 39.56 (10.99) | 47.18 (16.43)<sup>c</sup> | H = 5.83 | .054 |  |
| Delta TMT (B-A) | 15.39 (6.66)<sup>a</sup> | 19.20 (10.79) | 20.38 (13.60)<sup>c</sup> | F = .878 | .423 |  |
| Alexithymia (TAS-20) | 43.8 (8.1) | 36.9 (7.8) | 40.9 (6.4) | F = 3.33 | .045 | placebo > E2V |
| Trait emotion regulation (ERQ) |  |  |  |  |  |  |
| Cognitive reappraisal | 30.75 (3.84) | 29.06 (5.34) | 25.58 (5.20)<sup>d</sup> | F = 4.03 | .025 | placebo > pregnant |
| Emotional suppression | 11.44 (4.44) | 13.56 (3.50) | 11.54 (3.43) | F = 1.51 | .233 |  |
| Self-esteem (RSES) | 25.6 (3.8)<sup>a</sup> | 24.4 (4.3) | 23.5 (4.9)<sup>c</sup> | H = 1.88 | .391 |  |
| Anxiety (STAI) |  |  |  |  |  |  |
| Trait | 32.9 (5.5) | 37.0 (9.6) | 40.2 (11.5)<sup>c</sup> | F = 2.38 | .105 |  |
| State | 35.7 (6.3) | 38.1 (5.7) | 37.7 (5.7) | H = 2.83 | .243 |  |
| Affect ratings (ESR) |  |  |  |  |  |  |
| Anger | 1.00 (0.00) | 1.38 (0.81) | 1.47 (0.92) | H = 4.86 | .088 |  |
| Disgust | 1.00 (0.00) | 1.19 (0.54) | 1.27 (0.59) | H = 3.23 | .199 |  |
| Fear | 1.13 (0.34) | 1.13 (0.34) | 1.53 (1.06) | H = 1.76 | .414 |  |
| Sadness | 1.06 (0.25) | 1.13 (0.34) | 1.07 (0.26) | H = 0.49 | .784 |  |
| Happiness | 3.13 (0.34) | 2.81 (.98) | 2.73 (1.16) | F = 0.84 | .437 |  |
| Surprise | 1.53 (0.92) | 1.63 (0.72) | 2.2 (0.94) | H = 5.40 | .067 |  |
| Stress | 1.69 (0.95) | 2.31 (0.89) | 1.80 (1.01) | H = 2.85 | .241 |  |
| Depressive symptoms |  |  |  |  |  |  |
| BDI-2 | 6.2 (5.0)<sup>a</sup> | 8.0 (6.4) |  | U = 98.5 | .394 |  |
| EPDS |  |  | 5.9 (4.4)<sup>c</sup> |  |  |  |
| Sleep Quality (PSQI) Global | 5.00 (3.13)<sup>b</sup> | 6.29 (2.05)<sup>b</sup> | 6.43 (3.69)<sup>h</sup> | F = .91 | .413 |  |
| Sleep quality | .75 (.58) | 1.19 (.40) | 1.27 (.79)<sup>e</sup> | F = 3.36 | .045 | pregnant > placebo |
| Sleep latency | 1.50 (1.46) | 2.13 (1.78) | 1.64 (1.63)<sup>e</sup> | F = .67 | .535 |  |

|  |  |  |  |  |  |
| --- | --- | --- | --- | --- | --- |
| Sleep duration | .31 (.48) | .31 (.48) | .55 (.69)<sup>e</sup> | F = .77 | .472 |
| Sleep efficiency | .38 (.62) | .25 (.45) | .64 (1.03)<sup>e</sup> | F = 1.02 | .370 |
| Sleep disturbance | 1.00 (.39)<sup>b</sup> | 1.07 (.27)<sup>b</sup> | 1.29 (.49)<sup>h</sup> | F = 1.42 | .257 |
| Sleep medication use | 0.00 (0.00) | 0.00 (0.00) | 0.00 (0.00)<sup>e</sup> |  |  |
| Daytime dysfunction | .88 (6.2) | 1.13 (.81) | 1.18 (.60)<sup>e</sup> | F = .81 | .454 |
| Body Shape Perception (FFB) | 81.4 (24.4) | 78.6 (31.4) | 80.1 (30.7)<sup>c</sup> | H = .043 | .979 |
| Sexual Function (FSFI) |  |  |  |  |  |
| Desire | 6.3 (1.9) | 6.5 (1.9) | 6.0 (2.4)<sup>c</sup> | F = .21 | .810 |
| Arousal | 13.1 (3.7)<sup>b</sup> | 10.8 (3.3)<sup>f</sup> | 11.9 (3.8)<sup>d</sup> | F = 1.32 | .282 |
| Lubrication | 16.6 (1.2)<sup>b</sup> | 16.0 (.94)<sup>f</sup> | 16.2 (.94)<sup>d</sup> | F = 1.31 | .285 |
| Orgasm | 12.1 (1.8)<sup>b</sup> | 11.4 (1.3)<sup>f</sup> | 11.6 (1.6)<sup>d</sup> | F = .71 | .501 |
| Satisfaction | 9.0 (3.7)<sup>k</sup> | 7.0 (.71)<sup>k</sup> | 9.5 (4.4)<sup>i</sup> | F = .78 | .478 |
| Pain | 15.6 (3.5)<sup>g</sup> | 16.2 (1.7)<sup>j</sup> | 15.8 (2.0)<sup>d</sup> | F = .08 | .825 |
| Negative Emotional State Rating |  |  |  |  |  |
| View | -43.43 (28.06)<sup>a</sup> | -65.69 (33.37)<sup>a</sup> | -52.11 (40.33)<sup>b</sup> | F = 1.14 | .331 |
| Downregulation | -27.67 (41.06)<sup>a</sup> | -32.84 (30.56)<sup>a</sup> | -9.85 (61.36)<sup>b</sup> |  |  |
| Regulation Success | 15.76 (34.25)<sup>a</sup> | 32.85 (45.14)<sup>a</sup> | 42.26 (48.02)<sup>b</sup> | F = 1.33 | .248 |
| Positive Emotional State Rating |  |  |  |  |  |
| View | 55.43 (26.71)<sup>a</sup> | 71.41 (28.46) | 62.94 (35.56)<sup>c</sup> | F = 1.14 | .330 |
| Upregulation | 96.54 (31.65)<sup>a</sup> | 110.06 (25.40) | 99.00 (31.74)<sup>c</sup> |  |  |
| Regulation Success | 41.11 (22.97)<sup>a</sup> | 38.66 (12.95) | 36.06 (29.58)<sup>c</sup> | F = .22 | .895 |

*Note.* Data are the mean value with standard deviations in brackets.
Abbreviations: BDI, Beck-Depression-Inventory; E2, estradiol; E2V, estradiol valerate; EPDS, Edinburgh Postnatal Depression Scale; ERQ, Emotion Regulation Questionnaire; ESR, Emotional Self-Rating; FFB, Fragebogen zum Figurbewusstsein (Body Shape Questionnaire); FSFI, Female Sexual Function Index; PSQI, Pittsburgh Sleep Quality Index; RSES, Rosenberg self-esteem scale; STAI, State Trait Anxiety Index; TAS-20, Toronto Alexithymia Scale; TMT-A/B, Trail making test A/B; WST, Wortschatztest.
<sup>a</sup>n = 15, <sup>b</sup>n = 14, <sup>c</sup>n = 13, <sup>d</sup>n = 12, <sup>e</sup>n = 11 <sup>f</sup>n = 10, <sup>g</sup>n = 8, <sup>h</sup>n = 7, <sup>i</sup>n = 6, <sup>k</sup>n = 5.

#### Psychophysiological interaction (PPI) analysis

To investigate changes in functional connectivity of brain regions (physiological component) during the downregulation and view condition (psychological component) we conducted ROI-to-ROI PPI analyses (70) using the CONN Toolbox (71) Following Morawetz and colleagues (29), we performed a PPI analysis of (*1*) *the left amygdala with the left IFG*. Additionally, we explored connectivity of (*2*) *the left amygdala with the left MFG*. Time-series from these ROIs were determined, individual connectivity parameters for each regulation condition extracted and entered as dependent variables in separate univariate Analysis of Covariance (ANCOVA) with group (placebo, E2V, pregnant) as factor and covariates age and regulation success (29,30). To correct for multiple testing, the Bonferroni-corrected significance level was set to *p* < .025. Due to missing data, 14 females in the placebo, 15 in the E2V, and 14 in the pregnant group were included in the PPI analyses.

### Resting-state data analysis

Examining the same task-related connectivties at rest, ROI-to-ROI resting-state connectivity parameters of (*1*) *the left amygdala with the left IFG* as well as (*2*) *the left amygdala with the left MFG* were extracted using the CONN Toolbox (71). The effect of group (placebo, E2V, pregnant) on resting-state connectivity parameters was analyzed in an ANCOVA with age and regulation success score as covariates. To correct for multiple testing, the Bonferroni-corrected significance level was set to *p* < .025. Furthermore, we explored resting-state connectivity in (*3*) *an emotion downregulation network* consisting of previously reported ROIs (27,72; for detailed information see Supplement). For the resting-state analysis, data was missing for one pregnant participant.

### Regression analyses

First, separate linear regression analyses were conducted with brain activity during downregulation as predictor (left/right IFG; left/right MFG; left/right amygdala) of regulation success scores in each group, respectively. To correct for multiple testing related to laterality, the Bonferroni-corrected significance level was set to *p* < .025. Second, in each group, we conducted regression analyses of negative emotion regulation success as predictor of depression scores (EPDS for pregnant, BDI-2 for nonpregnant groups). Third, individual regression analyses were conducted with (1) bilateral amygdala activity during downregulation and (2) functional connectivity (left amygdala with left IFG; left amygdala with left MFG during downregulation) as predictors of depression scores. To correct for multiple testing the Bonferroni-corrected significance level was set to *p* < .025.

## Results

### Sample characteristics

Detailed sample characteristics are presented in Table1. Mean age between the groups was significantly different (H = 20.53, *p* < .001) with pregnant participants being older than nonpregnant participants. In terms of *cognitive abilities*, groups differed in processing speed (TMT-A: H = 8.92, *p* = .012), with pregnant females being slower than the placebo (*p* = .017) and E2V group (*p* = .004). However, groups showed no difference on the delta of TMT-B–A (*F*(2,43) = .88, *p* = .423), which is used to remove bias due to psychomotor functioning or visual sequencing (73). In terms of s*elf-report data*, the placebo and E2V group differed in alexithymia scores (TAS-20: *F*(2,46) = 3.33, *p* = .045, part-η^2^ = .137; placebo vs E2V group: *p* = .041) but no difference was found in comparison to the pregnant group (*p*s >. 478). Also, pregnant females reported reduced subjective sleep quality compared to females in the placebo group (*F*(2,40) = 3.36, *p* = .045; part-η^2^ = .144, placebo vs pregnant: *p* = .083), but groups did not differ on the total sleep quality index (*F*(2,34) = .91, *p* = .413). In terms of *hormones*, pregnant females had higher levels in all assessed sex hormones compared to both nonpregnant groups (*ps* < .031). The two nonpregnant groups only differed in E2 levels (*p* < .001) but not in progesterone and testosterone (*ps* > .451). In terms of *trait emotion regulation use* (ERQ), groups differed in cognitive reappraisal (*F*(2,43) = 4.03, *p* = .025, part-η^2^ = .164), with pregnant females reporting lower ratings compared to the placebo (*p* = .022) but not the E2V group (*p* = .194). E2V and placebo groups did not differ in their cognitive reappraisal ratings (*p* = .978; see Figure 2). No group differences appeared for emotional suppression (*F*(2,44) = 1.51, *p* = .233). For all other sample characteristics, no differences emerged (*ps* > .054).

**Figure 2.**
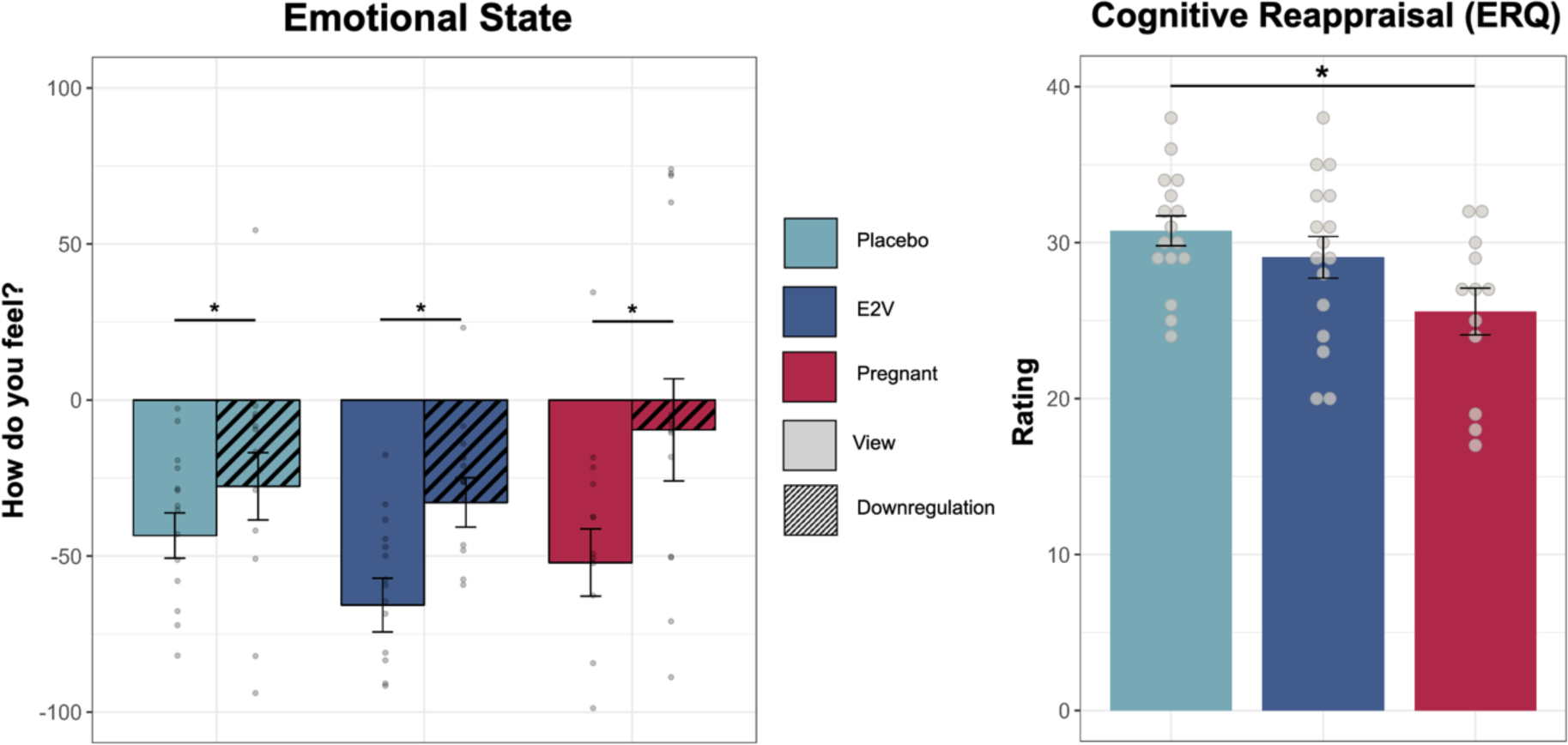
Emotional functions: Behavioral paradigm and self-report measures. *Note. Left:* Negative emotion regulation paradigm. Reduced negative emotional states were reported after the downregulation as compared to the view instruction across all groups. No significant differences between the groups emerged. *Right*: Cognitive reappraisal (ERQ); reduced self-report use of cognitive reappraisal in pregnant females compared to the placebo group. For statistical outcomes see Table 1. Data are the median (line) and interquartile range (box) per group with lower and upper quartile as error bars. \**p* < .05. Abbreviations: ERQ, Emotion Regulation Questionnaire.

### Behavioral results of negative emotion regulation

For negative emotional state ratings, a significant main effect of regulation was found (*F*(1,41) = 22.07, *p* < .001, part-η^2^ = .350), with reduced negative emotional state ratings after downregulation compared to view. The main effect of group was not significant (*F*(2,41) = 1.14, *p* = .331) and no significant interaction was revealed (*F*(2,41) = 1.44, *p* = .248). Likewise, groups did not differ significantly in regulation success (*F*(2,41) = 1.44*, p* = .248). See Table 1 and Figure 2 for details. Results for positive emotion regulation are reported in the Supplement and Table 1.

### Task-based fMRI results of emotion regulation

#### Whole-brain analysis

In a whole-brain approach, the 3×2 full factorial analysis showed a significant main effect of regulation, with an increased activation of the left MFG during downregulation (MNI (x,y,z) = -41, 7, 49 ; p_FWE_ < .001, k = 310; see Supplementary Table S1), while no significantly stronger activation during view was revealed (see Figure 3a).

**Figure 3.**
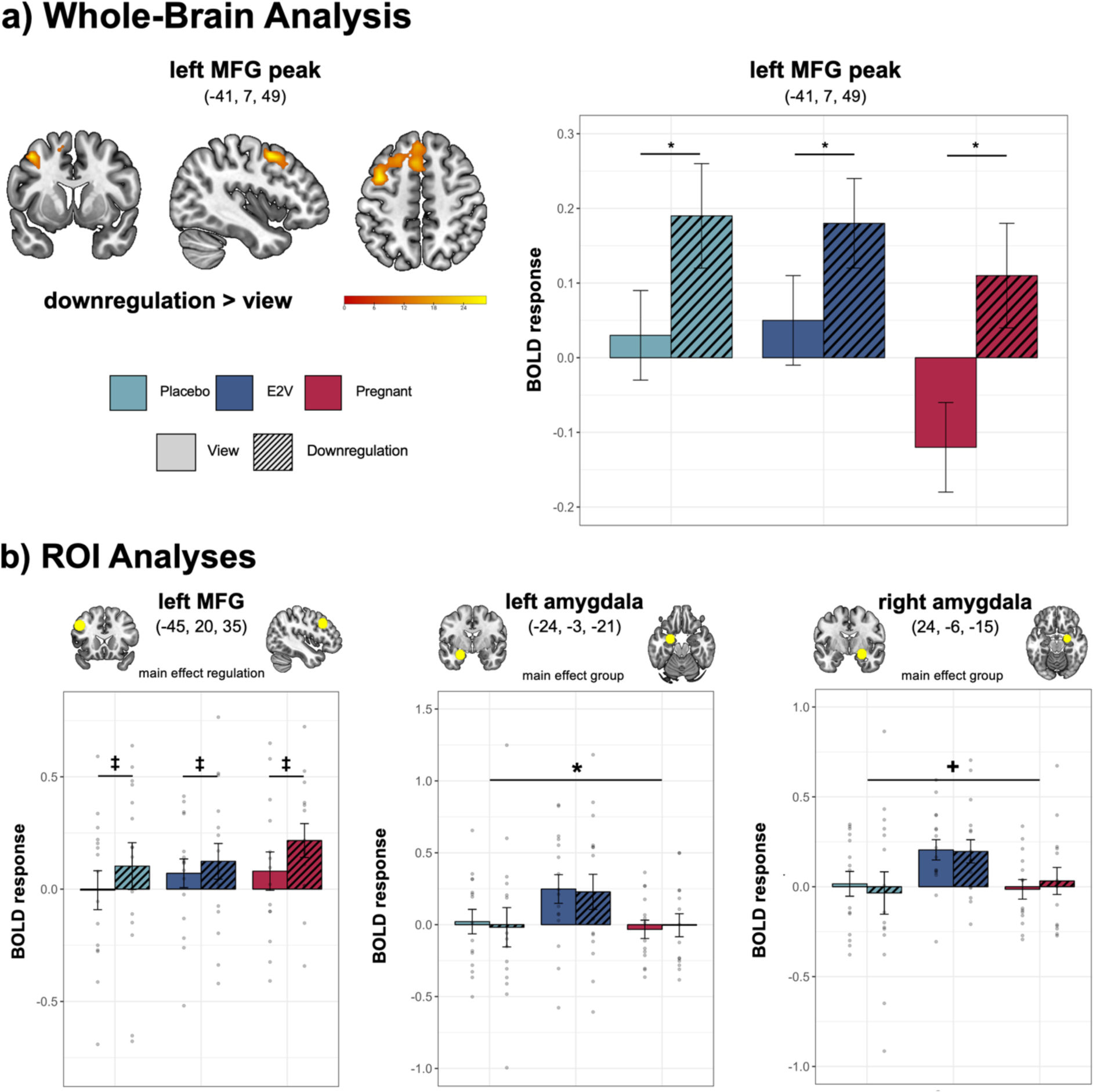
Whole-brain (a) and ROI analyses. **(b)** *Note.* a) Whole-brain analysis. Contrast downregulation > view; higher brain activation during downregulation was detected in the left MFG. The bar graph shows the mean activation per group and regulation condition with standard errors as error bars. b) ROI analyses. A tendency for a main effect of regulation was found for the left MFG with increased activity during downregulation compared to view; a significant main effect of group was found for the left amygdala and a tendency for a main effect of group found for the right amygdala. The bar graphs show the mean activation per group and regulation conditions with standard errors as error bars and individual data points. Coordinates are presented in MNI space. Results of the ANOVA effect: \**p* < .025, +*p* = .034, ‡ = .075. Abbreviations: MFG, middle frontal gyrus; ROI, region of interest.

#### Region-of-interest analyses

In line with whole-brain results, we observed a tendency for a main effect of regulation for the *left MFG* (*F*(1,40) = 3.33, *p* = .075, part-η^2^ = .077; see Figure 3b), with higher brain activity during downregulation compared to view (*p* = .003). No significant group effect (*F*(2,40) = .31, *p* = .736) nor interaction emerged (*F*(2,40) = 1.57, *p* = .220). For all other frontal ROIs (*right MFG, bilateral IFG*), no significant effects occurred (*ps* > .116).

For the *left amygdala*, a main effect of group emerged (*F*(2,40) = 4.71, *p* = .015,part-η^2^= .191; see Figure 3b), but no regulation effect (*F*(1,40) = .39, *p* = .538) nor interaction (*F*(2,40) = .02, *p* = .978). In terms of the group effect, E2V showed significantly higher activity compared to the pregnant group (*p* = .019), but not compared to the placebo group (*p* = .103). There was no difference between the pregnant and placebo group (*p* = .597). For the *right amygdala*, a tendency of a group effect corrected at Bonferroni-level emerged (*F*(2,40) = 3.67, *p* = .034, part-η^2^ = .155; see Figure 3b). No regulation effect (*F*(1,40) = .39, *p* = .538), nor interaction (*F*(2,40) = .29, *p* = .750) occurred. Post-hoc comparisons revealed a trend for higher activity in the E2V compared to the placebo (*p* = .057) but not compared to the pregnant group (*p* = .123). Activity in the placebo was not different to the pregnant group (*p =* 1.00).

#### PPI analysis

Controlling for age and regulation success, connectivity analysis of (*1*) *the left amygdala with the left IFG* revealed no significant differences during downregulation (*F*(2,38) = .47, *p* = .626) and view (*F*(2,38) = .10, *p* = .907). Likewise, groups did not differ in functional connectivity of (*2*) the *left amygdala with the left MFG* during downregulation (*F*(2,38) = 1.47, *p* = .243) and view (*F*(2,38) = .73, *p* = .487).

### Resting-state connectivity

No significant group differences emerged for (*1*) *the left amygdala with the left IFG* (*F*(2,41) = .58, *p* = .566), nor for (*2*) the *left amygdala with the left MFG* (*F*(2,41) = .65, *p* = .526), when controlling for age and regulation success. Resting-state connectivity within the emotion downregulation network also did not differ between groups (all *p_FDR_* > .357; detailed results provided in the Supplement and Supplementary Table S2).

### Correlation and regression

#### Brain activity and regulation success

In pregnant females, *left amygdala* activity during downregulation was significantly negatively correlated with regulation success (r = -.60, *p* = .012; see Figure 4a). Also, the regression model was significant, with *left amygdala* activity during downregulation predicting regulation success (Model: R^2^ = .36, *F*(1,12) = 6.76, *p* = .023; activity: beta = -.60, *t*(13) = -2.60, *p* = .023). Brain activity during downregulation for all other ROIs was not associated with regulation success in the pregnant group (*p*s > .140). In both nonpregnant groups, no significant associations of brain activity during downregulation and regulation success emerged (*p*s > .100).

**Figure 4.**
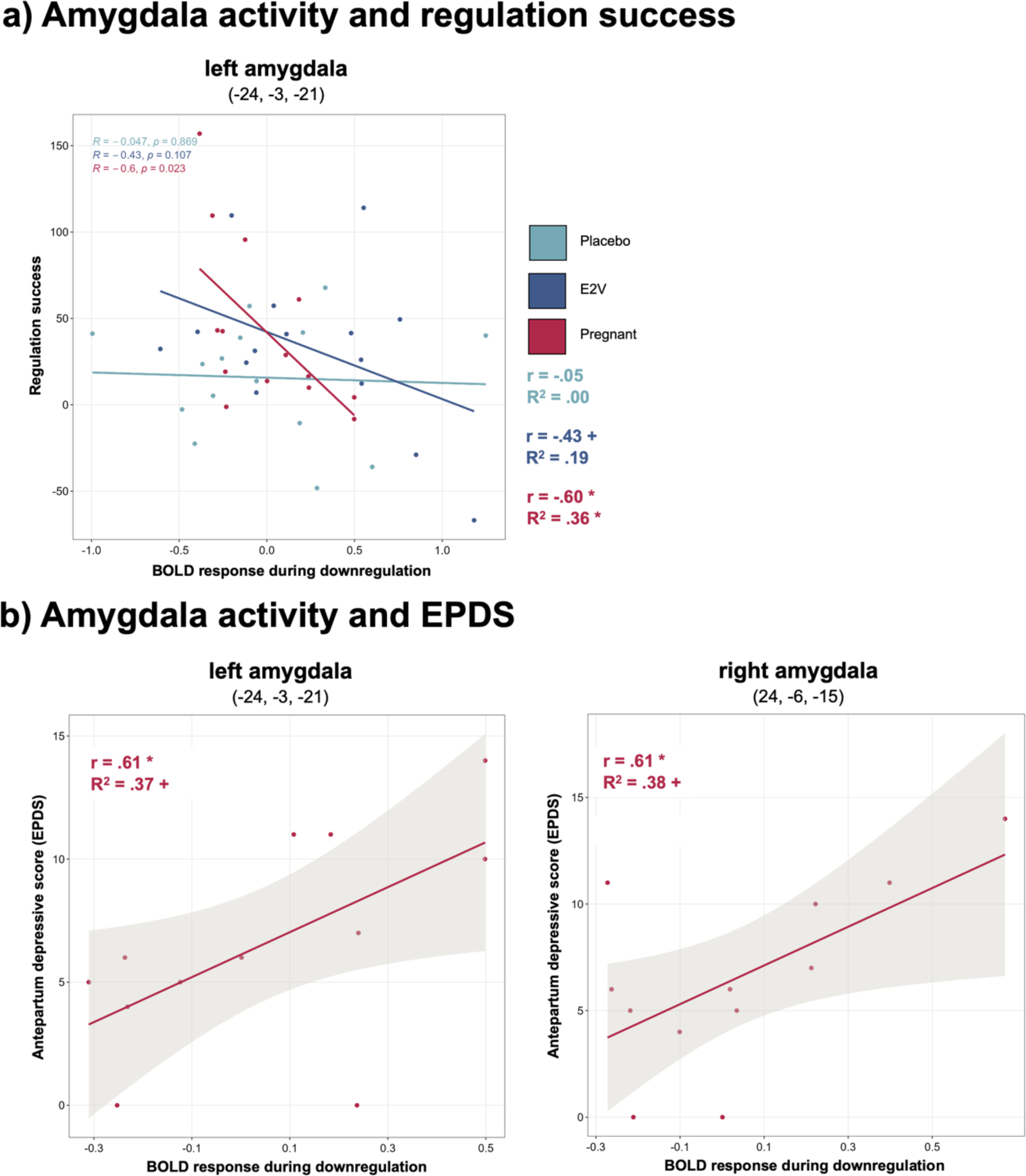
Relations of amygdala activity during downregulation, negative emotion regulation success, and antepartum depressive scores (EPDS) Note. a) Amygdala activity and regulation success. Increased left amygdala activity during downregulation significantly predicts reduced regulation success in pregnant females. b) Amygdala activity and antepartum depressive scores (EPDS) in pregnant females. *Left*: Increased activity in the left amygdala during downregulation was significantly related to higher EPDS scores. *Right*: Increased activity in the right amygdala during downregulation was significantly associated to higher EPDS scores. Coordinates are presented in MNI space. \**p* < .025, +*p* < .05. Abbreviations: EPDS, Edinburgh Postnatal Depression Scale.

#### Regulation success and depression scores

Across all groups, no significant associations of regulation success on depression scores occurred (*ps* > .121).

#### Brain activity and depression scores

In pregnant females, significant associations between bilateral amygdala reactivity and depression scores (EPDS) emerged (*left*: r = .61, *p* = .018; *right*: r = .61, *p* = .017; see Figure 4b). With a Bonferroni-corrected significance-level, the regression model showed borderline significance on EPDS scores for both the *left amygdala* (Model: R^2^ = .37, *F*(1,10) = 5.89, *p* = .036; beta = .61, *t*(11) = 2.43, *p* = .036) and the *right amygdala* during downregulation (Model: R^2^ = .38, *F*(1,10) = 6.01, *p* = .034; beta = .61, *t*(11) = 2.45, *p* = .034). Further, connectivity parameters of the *left IFG with left amygdala* showed a tendency for a positive correlation with EPDS scores (r = .439, *p* = .077), but connectivity of the *left MFG with left amygdala* showed no correlation with EPDS scores (r = .242, *p* = .224). In the nonpregnant groups, no significant associations of amygdala activity or functional connectivity during downregulation on depression scores (BDI-2) emerged (*ps* > .107). Detailed results are provided in Table 2.

**Table 2.**
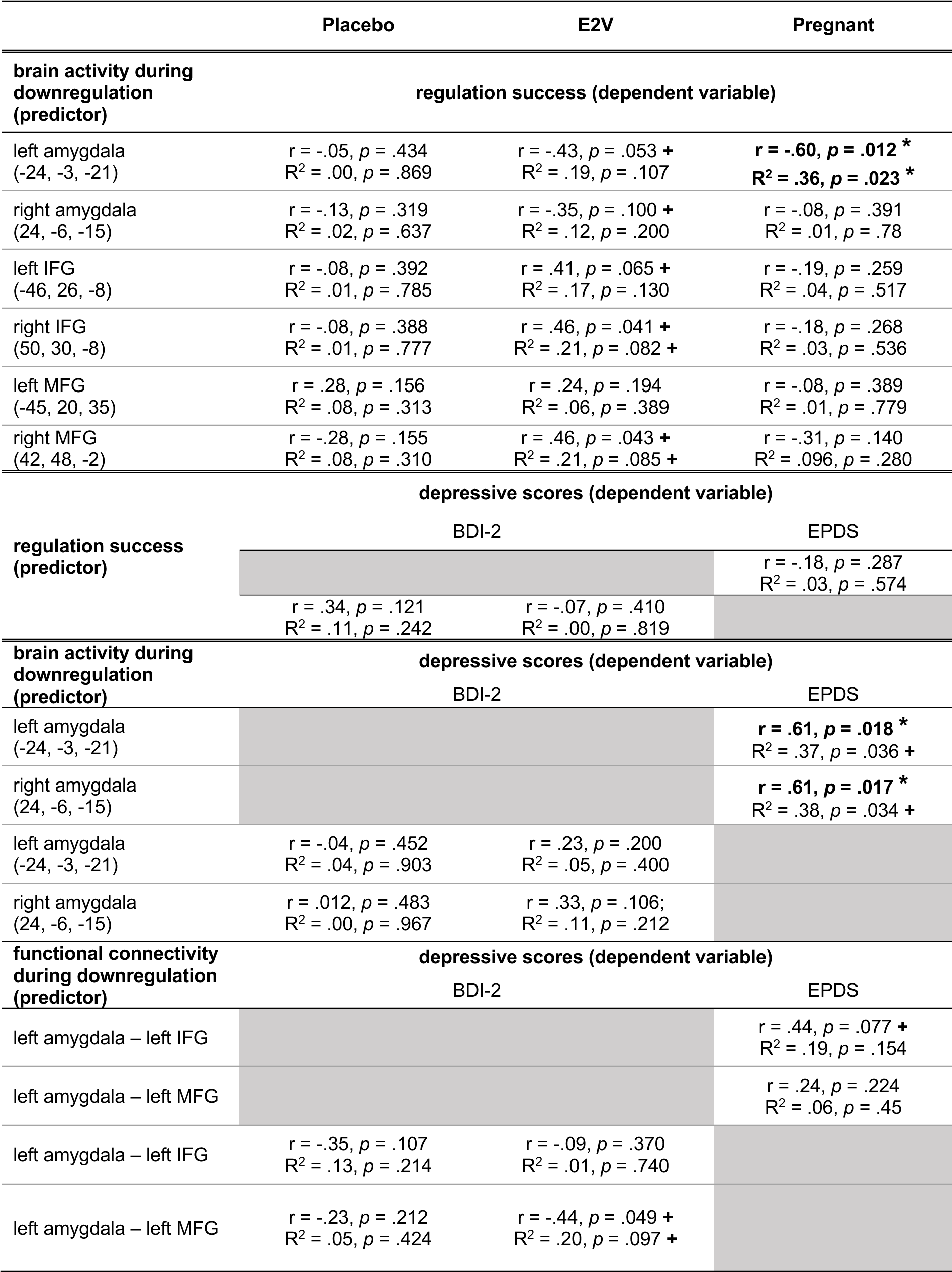
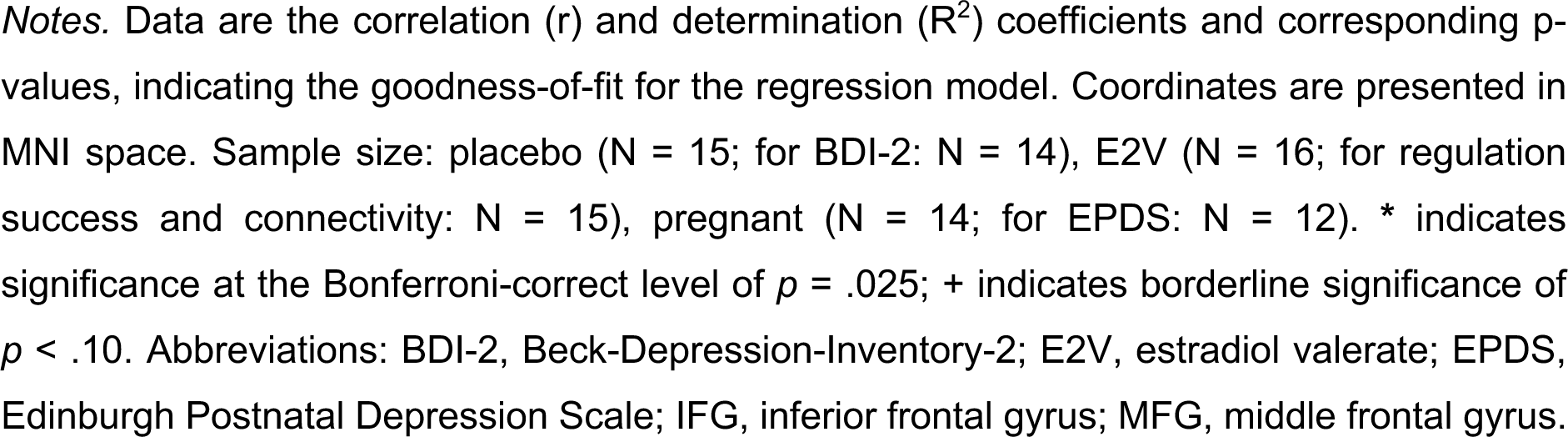
Relations of brain activity, emotion regulation success, and depressive scores across groups.

|  | Placebo | E2V | Pregnant |
| --- | --- | --- | --- |
| **brain activity during downregulation (predictor)** | **regulation success (dependent variable)** |  |  |
| left amygdala (-24, -3, -21) | $r = -.05, p = .434$<br>$R^2 = .00, p = .869$ | $r = -.43, p = .053 +$<br>$R^2 = .19, p = .107$ | $r = -.60, p = .012 *$<br>$R^2 = .36, p = .023 *$ |
| right amygdala (24, -6, -15) | $r = -.13, p = .319$<br>$R^2 = .02, p = .637$ | $r = -.35, p = .100 +$<br>$R^2 = .12, p = .200$ | $r = -.08, p = .391$<br>$R^2 = .01, p = .78$ |
| left IFG (-46, 26, -8) | $r = -.08, p = .392$<br>$R^2 = .01, p = .785$ | $r = .41, p = .065 +$<br>$R^2 = .17, p = .130$ | $r = -.19, p = .259$<br>$R^2 = .04, p = .517$ |
| right IFG (50, 30, -8) | $r = -.08, p = .388$<br>$R^2 = .01, p = .777$ | $r = .46, p = .041 +$<br>$R^2 = .21, p = .082 +$ | $r = -.18, p = .268$<br>$R^2 = .03, p = .536$ |
| left MFG (-45, 20, 35) | $r = .28, p = .156$<br>$R^2 = .08, p = .313$ | $r = .24, p = .194$<br>$R^2 = .06, p = .389$ | $r = -.08, p = .389$<br>$R^2 = .01, p = .779$ |
| right MFG (42, 48, -2) | $r = -.28, p = .155$<br>$R^2 = .08, p = .310$ | $r = .46, p = .043 +$<br>$R^2 = .21, p = .085 +$ | $r = -.31, p = .140$<br>$R^2 = .096, p = .280$ |
|  | **depressive scores (dependent variable)** |  |  |
| **regulation success (predictor)** | BDI-2 |  | EPDS |
| | | | $r = -.18, p = .287$<br>$R^2 = .03, p = .574$ |
| | $r = .34, p = .121$<br>$R^2 = .11, p = .242$ | $r = -.07, p = .410$<br>$R^2 = .00, p = .819$ | |
| **brain activity during downregulation (predictor)** | **depressive scores (dependent variable)** |  |  |
|  | BDI-2 |  | EPDS |
| left amygdala (-24, -3, -21) | | | $r = .61, p = .018 *$<br>$R^2 = .37, p = .036 +$ |
| right amygdala (24, -6, -15) | | | $r = .61, p = .017 *$<br>$R^2 = .38, p = .034 +$ |
| left amygdala (-24, -3, -21) | $r = -.04, p = .452$<br>$R^2 = .04, p = .903$ | $r = .23, p = .200$<br>$R^2 = .05, p = .400$ | |
| right amygdala (24, -6, -15) | $r = .012, p = .483$<br>$R^2 = .00, p = .967$ | $r = .33, p = .106$<br>$R^2 = .11, p = .212$ | |
| **functional connectivity during downregulation (predictor)** | **depressive scores (dependent variable)** |  |  |
|  | BDI-2 |  | EPDS |
| left amygdala – left IFG | | | $r = .44, p = .077 +$<br>$R^2 = .19, p = .154$ |
| left amygdala – left MFG | | | $r = .24, p = .224$<br>$R^2 = .06, p = .45$ |
| left amygdala – left IFG | $r = -.35, p = .107$<br>$R^2 = .13, p = .214$ | $r = -.09, p = .370$<br>$R^2 = .01, p = .740$ | |
| left amygdala – left MFG | $r = -.23, p = .212$<br>$R^2 = .05, p = .424$ | $r = -.44, p = .049 +$<br>$R^2 = .20, p = .097 +$ | |

*Notes.* Data are the correlation ( $r$ ) and determination ( $R^2$ ) coefficients and corresponding $p$ -values, indicating the goodness-of-fit for the regression model. Coordinates are presented in MNI space. Sample size: placebo ( $N = 15$ ; for BDI-2: $N = 14$ ), E2V ( $N = 16$ ; for regulation success and connectivity: $N = 15$ ), pregnant ( $N = 14$ ; for EPDS: $N = 12$ ). \* indicates significance at the Bonferroni-correct level of $p = .025$ ; + indicates borderline significance of $p < .10$ . Abbreviations: BDI-2, Beck-Depression-Inventory-2; E2V, estradiol valerate; EPDS, Edinburgh Postnatal Depression Scale; IFG, inferior frontal gyrus; MFG, middle frontal gyrus.

## Discussion

For the first time, behavioral and neural emotion regulation were investigated in healthy first-time pregnant females in the second trimester and compared to nulliparous females naturally-cycling (NC) females in the early follicular phase, who either received E2 valerate to increase E2 levels (E2V group) or a placebo to maintain low E2 levels (placebo group). While pregnant females reported the lowest tendency to use cognitive reappraisal during every-day life, we did not observe significant group differences in emotion regulation performance. This was also mirrored in frontal activity and functional connectivity, where pregnant females showed no differences compared to the nonpregnant groups. However, amygdala reactivity differed between groups. In pregnant females, amygdala activity during downregulation predicted reduced regulation success and related to increased self-reported depression symptoms. Thus, present results contribute several insights to the limited literature on emotional functions and neural activity during pregnancy, which will be discussed in the following.

Contrary to our expectations, *pregnant females in the second trimester showed no difference in whole-brain fMRI activity compared to nonpregnant females*. While considerable changes in brain structure and function related to pregnancy have been reported, most of this evidence in humans stems from pre-vs post-pregnancy comparisons or cross-sectional studies in pregnant vs nonpregnant women (3,10). Up until now, the only neuroimaging studies during antepartum used scalp-recorded near-infrared spectroscopy (NIRS; 18) and electro-encephalography (EEG; 17). While NIRS showed different prefrontal activation in response to emotional stimuli between trimesters of pregnancy, no differences compared to nonpregnant controls were reported (18). Using EEG, distinctions in neural recruitment during emotional processing were recorded for third trimester pregnant compared to nonpregnant women (17). Little is known about when and how reported neural changes occur in human pregnancy and if the second trimester could be a phase in which functional differences are not yet apparent using fMRI. For example, whereas MRI from *pre-pregnancy to postpartum* revealed reduced gray matter volume in anterior and posterior midline structures, bilateral temporal- and prefrontal cortex, including IFG, compared to nulliparous women (13,14), MRI in primiparous women in the *second trimester* showed smaller gray matter volume merely in the left putamen (12). Research in female rodents supports that processes underlying pregnancy-related neural changes are especially prominent during late pregnancy and throughout the postpartum period, among others due to the involved steroid hormones (74,75). Therefore, it may not be until the third trimester of pregnancy, when hormone levels have increased even more, or until postpartum, when hormone levels dropped sharply, that functional differences can be observed in whole-brain fMRI.

Yet, ROI-analyses indicated hypothesized group differences in the left amygdala as well as borderline significant differences in the right amygdala. Here the E2V group showed higher activity, regardless of regulation condition, compared to the pregnant in the left amygdala and a trend for the placebo group in the right amygdala. As illustrated in Figure3, the observed *activity pattern in the amygdala shows an inverted-U shape response in relation to E2 levels*. This reflects an activation pattern with the highest values in the medium range and low levels at the extremes. This was already observed in NC women, for which activity in the medial posterior hippocampus was reduced under low and supraphysiological E2 levels, whereas it was enhanced for physiologically high E2 levels (47). Characteristics of different estrogen receptors and their expression ratios are proposed to underlie these E2-dependent neuronal patterns (6,47,76). Thus, our data suggests that the inverted-U-shaped E2 dose-response relation might be applied to the amygdala region in response to emotional stimuli. However, E2 is not the only ovarian hormone involved in pregnancy with an influence on amygdala function and it should be carefully examined whether the observed differences can solely be attributed to E2-specific effects. For example, progesterone and testosterone are suggested to modulate amygdala activity in response to emotional stimuli in NC women and also increase and fluctuate during peripartum (3,6,77–79). Although it is not yet clear how the dynamic and complex hormonal milieu during pregnancy influences brain function, research in rodents shows that several pregnancy-related hormones have an influence on neural excitability (75), including the amygdala (74). Eventually these hormonal changes facilitate the development of maternal behavior (74,75), which was supported in human mothers, particularly in response to infant cues (80,81). Apart from maternal functions, the amygdala is generally involved in socioemotional processing as well as in identifying and appropriately responding to salient environmental stimuli (82–84). Hence, as part of an “maternal caregiving circuit” (85), it is apparent and adaptive that pregnancy-related plasticity processes affect the amygdala and thus influence emotional functions – all of which promotes maternal behavior (74). As the present study provides the first data on amygdala function in human pregnancy, associations with regulation success and mood symptoms were specifically assessed.

Remarkably, *in the pregnant group left amygdala activity during downregulation significantly predicts negative emotion regulation success,* whereby pregnant females with increased amygdala activity were less successful in regulating their emotional state. This is in line with evidence of reduced amygdala activity when emotions have been regulated successfully in men and nonpregnant women (29,32), as well as with evidence of enhanced amygdala activation when individuals with impaired regulation abilities engage in cognitive reappraisal (45). Thus, the same neural mechanism underlying successful emotion regulation seems to apply to our sample of pregnant females. At the same time our results suggest that the association of amygdala activity and regulation success could be more pronounced for pregnant women, possibly due to extreme E2 levels: while the amygdala x regulation success association was significant in the pregnant group, only a tendency emerged in the E2V group, and no significant relation occurred in the placebo group. Specifically, the heightened amygdala activity in pregnant females could indicate increased sensitivity or susceptibility (20), which, in relation to reduced regulation success, proposes a risk marker for mental ill-health. Amygdala hyperresponsiveness is associated with lower resilience, i.e., reduced capacity to tolerate stress, heightened vulnerability for depression, and has been proposed as a transdiagnostic factor for psychopathologies (86). For example, increased amygdala activity for negative stimuli is seen in nonpregnant patients with depression as compared to healthy controls (87–91). Evidence on neural correlates of peripartum depression is still scarce and inconclusive, but data in our *pregnant sample indicated a positive correlation of bilateral amygdala activity during downregulation and depression scores (EPDS)*. This was supported by the regression of amygdala activity predicting depressive scores, for which borderline significance was reached. Besides, in our pregnant group *a trend for a positive correlation between functional connectivity of the left IFG with the left amygdala during downregulation and EPDS scores* emerged. Although we cannot draw conclusions on effective connectivity, research supports that frontal regions are recruited to reduce amygdala activation during successful cognitive reappraisal (29). Thereby, our findings could reflect that women who need to exert more neural regulatory effort to reduce amygdala activity and regulate their emotions successfully also have higher depression scores. The relation of *regulation success and depressive scores was not found significant in our group of healthy pregnant females*. Yet, an association of emotion regulation abilities with depression scores has been reported for females diagnosed with PPD before (1,41). Presumably, the healthy status of our pregnant sample might account for these null findings, at least at the timepoint of their second trimester. Higher mood instability has been reported from the third trimester onwards until the early postpartum (19), so that changes in depression scores, regulation success as well as their relation in healthy pregnant women might only be observed at a later stage. Nevertheless, the present study is the first to show that amygdala activity in response to emotional stimuli is associated with early mood symptoms in the second trimester of pregnancy. Reduced regulation success is a risk marker for peripartum depression and anxiety (1,25,41,42), introducing heightened amygdala activity during pregnancy as a neural vulnerability correlate for peripartum depression. To verify these associations, future research should assess amygdala response ideally before, during and after pregnancy and monitor symptoms of depression, both in mothers with and without peripartum depression.

Comparing the groups on emotion regulation performance*, pregnant females were just as successful as nonpregnant females in regulating their emotional state* when confronted with emotional stimuli and asked to apply cognitive reappraisal. While this confirms and extends results in nulliparous women with low vs high E2 levels, who showed no difference in behavioral emotion regulation success (33,34), this contradicts reports of altered emotional functions among pregnant women (2,16–18). Inconsistent findings could be explained by the timepoint of our assessment, which was conducted in the *second trimester* of pregnancy. Previous studies, however, reported increased mood instability, greater emotional sensitivity, and altered evaluation of emotional stimuli during the *third trimester* (16,17,19). Pregnancy and motherhood mark transition phases which are accompanied by several psychosocial challenges, including diverse and varying internal experiences and societal expectations (3). Over the course of pregnancy, these challenges can change and ultimately influence emotion regulation abilities and the use of (mal)adaptive regulation strategies (3,92). Consequently, emotion regulation should be investigated in different peripartum stages to draw conclusions about potential changes in regulation abilities and the use of regulation strategies throughout pregnancy. The absence of behavioral differences in the present sample was mirrored on the neural level, where *no group differences were found for functional activity related to emotion regulation in whole-brain- and frontal ROI analyses*. This was contrary to our hypothesis based on pregnancy-related plasticity in brain regions associated to emotion regulation (13,14). Instead, in accordance with the emotion regulation literature*, all participants had increased activity in the left MFG during the downregulation of negative emotions* (21,26,27), indicating the same neural mechanism of emotion regulation being recruited during pregnancy. Notably, however, for the pregnant group we observed pronounced activation differences in the left MFG during the downregulation compared to the view condition. While this observation must be interpreted carefully, the activation differences during cognitive reappraisal could indicate higher regulation effort exerted by pregnant females. This has been suggested for nonpregnant women, in which increased frontal activation during emotion regulation, i.e., neural effort, was found in women with low compared to high E2 levels (33). Replication using fMRI in a larger sample of antepartum women might reveal whether increased left MFG activation during downregulation can be verified. Further, *we did not find any group differences in functional connectivity during downregulation, nor in resting-state connectivity.* Hence, any neural plasticity in regions and circuits involved in emotion regulation (3,13,14) did not seem to affect neural downregulation of negative emotions in healthy pregnant females during the second trimester. However, parity is proposed to modulate both neural activity and connectivity (20), indicating that differences might be observed in multiparous as compared to primiparous and nulliparous women. Insufficient difficulty of the functional paradigm could also explain lack of group differences, that is, recording ceiling effects instead of detecting differences in regulation abilities. Yet, the present paradigm is an established fMRI paradigm and was used in previous studies (29,31,67).

Notably, *pregnant females reported the lowest tendency of using cognitive reappraisal in their daily life*. Previous evidence on higher mood instability in pregnant women has also been based on self-reports (19). Presumably, rather than failing to regulate emotions, pregnant women could perceive reduced emotion regulation abilities or experience more fluctuating emotions to regulate. Possibly, they might also rely on other strategies than cognitive reappraisal in daily life, such as interpersonal regulation strategies (19,41). Since the present self-report questionnaire only asked about cognitive reappraisal and expressive suppression, future studies should assess the use of various strategies.

Our data have some limitations that may influence data interpretation and raise ideas for future research. Despite assessing human primiparous females in the MRI-scanner, which poses some challenges, we acknowledge the small sample size that should be taken into consideration when interpretating the data. Even though our study reported large effect sizes, the small sample resulted in low power. Our cross-sectional study only included healthy primiparous females during the second trimester of pregnancy. To understand pregnancy-related changes in emotional functions and neuronal plasticity, future research should conduct repeated-measures designs in different stages of pregnancy and the postpartum period. Cross-sectional designs comparing primiparous and multiparous mothers would improve our understanding of the potential modulating role of parity on the brain (20). Besides replication in healthy samples, it is of utmost importance to assess clinical samples of ante- and postpartum women regarding their behavioral and neural emotion regulation to characterize and confirm the outlined risk associations.

The results of the present study have implications not only in providing first insights of fMRI during human pregnancy and highlighting a risk association of amygdala function, regulation success, and depression scores in pregnant females, but also may have clinical implications. Up to 80% of pregnant women with high anxiety or depressive symptoms go unidentified and do not receive treatment, which has long-term consequences for mother and child (87). Recognizing the peripartum period as a unique opportunity to identify women at risk for pregnancy-related mental health problems (10), the results of the present study suggest that vulnerability markers and risk associations could be observed in antepartum self-report, behavior, and neural function. Concretely, screening emotion regulation abilities and assessing mood symptoms during and after pregnancy and subsequently promoting adaptive emotion regulation should be considered as preventive intervention for PPD (1,3). As a resilience factor for mental health, fostering adaptive regulation strategies, like cognitive reappraisal, not only affects maternal wellbeing but also the health and development of the child as well as parent-child relations (39–41).

## Data Availability

Datasets used in the present study are available upon reasonable request from the corresponding authors.

## Author Contribution

FW: Formal analysis; Investigation; Visualization; Writing – original draft. LK: Supervision; Methodology; Validation; Writing – review & editing. ER: Data curation; Writing – review & editing. CM: Methodology; Validation; Writing – review & editing. ISP: Conceptualization; Funding acquisition; Methodology; Writing – review & editing. AS: Supervision; Writing – review & editing. BD: Conceptualization; Funding acquisition; Methodology; Project administration; Resources; Supervision; Validation; Writing – review & editing.

## Notes

**Conflict of Interest Disclosure.** The authors declare no competing interests.

### Competing Interest Statement

The authors have declared no competing interest.

### Clinical Trial

NCT06312033

### Funding Statement

This study was funded by the Center for Integrative Neuroscience, Tuebingen, Germany (CIN; EXC307, MRTG Pregnancy & the Brain) and the International Research Training Group "Women's Mental Health across the Reproductive Years" (IRTG2804).

### Author Declarations

Ethics Committee of the Medical Faculty of the University of Tuebingen gave ethical approval for this work.

