## Supplementary Information for "Neural emotion regulation during pregnancy – a fMRI study investigating a transdiagnostic mental health factor in healthy first-time pregnant women"

### Positive emotion regulation paradigm

The positive emotion regulation paradigm outside the MRI-scanner followed the same design and procedure as the negative emotion regulation paradigm, but the participants were exposed to 24 positively valanced pictures. In an 1) *upregulation* instruction, they were asked to increase their emotional response toward the picture (indicated by an upward pointing arrow) or 2) to experience arising emotions towards the picture without changing them in a *view* instruction (indicated by an equal sign). Each picture was shown twice for the duration of 8sec with the instruction for upregulation or view preceding for 2sec. After each picture, participants rated their emotional state on a continuous visual rating scale from very negative to very positive. Emotional state ratings acquired per picture and instruction were summarized in mean values for the view and upregulation condition for each participant. To compare between groups and conditions, mixed between-within-subjects ANOVAs were performed with mean state ratings as a dependent interval variable for positive emotions. Group (placebo, E2V, pregnant) was the between-subjects factor and regulation (view, upregulation) was the within-subject factor. In case of significant main or interaction effects, Bonferroni-adjusted post-hoc tests were performed to assess and interpret mean differences on emotional state ratings. To assess emotion regulation success, we subtracted mean emotional state ratings for view from mean ratings for the upregulation condition for each participant. For the emotional state ratings, data was missing for three females (one placebo, two pregnant).

The main effect for positive regulation was significant ($F(1,41) = 130.81$, $p < .001$, part-$\eta^2 = .761$), with enhanced emotional state ratings after upregulation compared to view. The main effect of group was not significant ($F(2,41) = 1.14$, $p = .330$). No significant interaction between group and regulation was found ($F(2,41) = -1.78$, $p = .837$). The groups also did not differ in emotion regulation success of positive emotions ($H = .221$, $p = .895$). See Table 1 in the main Manuscript for details.

**Resting-state data analysis: emotion downregulation network**

Resting-state connectivity was examined in an emotion downregulation network, which consisted of the following, previously reported ROIs (1,2): left IFG (MNI(x,y,z): -46, 26, -8), right IFG (MNI(x,y,z): 50, 30, -8), right MFG (MNI(x,y,z): 41, 23, 41), left superior frontal gyrus (SFG; MNI(x,y,z): -9, 11, 62), left middle temporal gyrus (MTG; MNI(x,y,z): -60, -38, -2), left superior temporal gyrus (STG; MNI(x,y,z): -42, -56, 24), and right supramarginal gyrus (SMG; MNI(x,y,z): 58, -54, 38). These ROIs were defined with the MarsBarR toolbox (3; 5mm sphere around the reported MNI peaks) and only used for the resting-state analysis. Using the CONN Toolbox (4), time-series from these ROIs were determined and individual ROI-to-ROI connectivity parameters extracted. Using IBM SPSS Statistics (version 27.0), we analyzed the effect of the factor group (placebo, E2V, pregnant) on resting-state connectivity parameters in an ANCOVA with age and negative emotion regulation success score as covariates. We performed separate analyses for each ROI-to-ROI resting-state connectivity ($m = 21$ comparisons) and corrected for multiple testing by applying the false discovery rate (FDR)-controlling Benjamini-Hochberg procedure with an FDR of $q < .05$. For the resting-state analysis, data was missing for one pregnant female.

After FDR-correction, the ANCOVA with covariates age and regulation success indicated no significant difference between the groups for any of the resting-state connectivities within the downregulation network (all $p_{FDR} > .357$). Detailed results are provided in Supplementary Table S2.

**Table S1.**

*Activation during emotion downregulation compared to view: activation cluster including peaks.*

| Contrast | #Cluster | *k* | Brain region | MNI coordinates | | | z-score |
|---|---|---|---|---|---|---|---|
| | | | | x | y | z | |
| downregulation > view | 1 | 315 | Middle frontal gyrus | -41 | 7 | 49 | 4.85 |
| | | | Supplementary motor area | -7 | 21 | 49 | 4.30 |
| | | | Medial frontal gyrus | 7 | 51 | 39 | 4.30 |

*Notes.* Peaks are identified with MNI coordinates. Significance level of $p < 0.001$, corrected for multiple comparisons with cluster-wise correction. K, cluster size.

**Table S2.**

*Resting-state connectivity within emotion downregulation network between groups*

| ROI: | left IFG | right IFG | right MFG | left SFG | left MTG | left STG | right SMG |
|---|---|---|---|---|---|---|---|
| **left IFG**<br>(-46, 26, -8) | | | | | | | |
| **right IFG**<br>(50, 30, -8) | $F$ = .93,<br>$p_{uncorr}$ = .404<br>$p_{FDR}$ = 1.06 | | | | | | |
| **right MFG**<br>(41, 23, 41) | $F$ = 2.07,<br>$p_{uncorr}$ = .140<br>$p_{FDR}$ = .735 | $F$ = .18<br>$p_{uncorr}$ = .838<br>$p_{FDR}$ = .926 | | | | | |
| **left SFG**<br>(-9, 11, 62) | $F$ = 2.74<br>$p_{uncorr}$ = .077<br>$p_{FDR}$ = .539 | $F$ = .38,<br>$p_{uncorr}$ = .688<br>$p_{FDR}$ = .963 | $F$ = 0.41,<br>$p_{uncorr}$ = .669<br>$p_{FDR}$ = 1.00 | | | | |
| **left MTG**<br>(-60, -38, -2) | $F$ = .19<br>$p_{uncorr}$ = .829<br>$p_{FDR}$ = .967 | $F$ = 1.30,<br>$p_{uncorr}$ = .285<br>$p_{FDR}$ = .855 | $F$ = 0.34,<br>$p_{uncorr}$ = .714<br>$p_{FDR}$ = .937 | $F$ = 1.87,<br>$p_{uncorr}$ = .168<br>$p_{FDR}$ = .588 | | | |
| **left STG**<br>(-42, -56, 24) | $F$ = 3.78<br>$p_{uncorr}$ = .032<br>$p_{FDR}$ = .672 | $F$ = 0.24,<br>$p_{uncorr}$ = .787<br>$p_{FDR}$ = .972 | $F$ = 0.55,<br>$p_{uncorr}$ = .581<br>$p_{FDR}$ = 1.02 | $F$ = 0.11,<br>$p_{uncorr}$ = .895<br>$p_{FDR}$ = .940 | $F$ = 0.02,<br>$p_{uncorr}$ = .981<br>$p_{FDR}$ = .981 | | |
| **right SMG**<br>(58, -54, 39) | $F$ = 3.69<br>$p_{uncorr}$ = .034<br>$p_{FDR}$ = .357 | $F$ = 0.52,<br>$p_{uncorr}$ = .600<br>$p_{FDR}$ = .969 | $F$ = 0.89,<br>$p_{uncorr}$ = .421<br>$p_{FDR}$ = 982 | $F$ = 0.63,<br>$p_{uncorr}$ = .540<br>$p_{FDR}$ = 1.03 | $F$ = 1.93,<br>$p_{uncorr}$ = .159<br>$p_{FDR}$ = .668 | $F$ = 0.78,<br>$p_{uncorr}$ = .467<br>$p_{FDR}$ = .981 | |

*Notes.* F-statistic (2,38) of ANCOVA (dependent variable: connectivity parameter; factor: group; covariates: age, regulation success) with uncorrected and FDR-corrected p-values. No significant results were found at the Benjamini-Hochberg corrected FDR of $q < .05$. Coordinates are presented in MNI space. Sample size: placebo (N = 16), E2 valerate (N = 16), pregnant (N = 14). Abbreviations: IFG, inferior frontal gyrus; MFG, middle frontal gyrus; SFG, superior frontal gyrus; SMG, supramarginal gyrus; STG, superior temporal gyrus.
